# Flattening the curve is not enough, we need to squash it: An explainer using a simple model

**DOI:** 10.1101/2020.03.30.20048009

**Authors:** ES McBryde, MT Meehan, JM Trauer

**Affiliations:** Australian Institute of Tropical Health and Medicine, James Cook University; School of Public Health and Preventive Medicine, Monash University

## Abstract

**Background:** Around the world there are examples of both effective control (e.g., South Korea, Japan) and less successful control (e.g., Italy, Spain, United States) of COVID-19 with dramatic differences in the consequent epidemic curves. Models agree that flattening the curve without controlling the epidemic completely is insufficient and will lead to an overwhelmed health service. A recent model, calibrated for the UK and US, demonstrated this starkly.

**Methods:** We used a simple compartmental deterministic model of COVID-19 transmission in Australia, to illustrate the dynamics resulting from shifting or flattening the curve versus completely squashing it.

**Results:** We find that when the reproduction number is close to one, a small decrease in transmission leads to a large reduction in burden (i.e., cases, deaths and hospitalisations), but achieving this early in the epidemic through social distancing interventions also implies that the community will not reach herd immunity.

**Conclusions:** Australia needs not just to shift and flatten the curve, but to squash it by getting the reproduction number below one. This will require Australia to achieve transmission rates at least two thirds lower than those seen in the most severely affected countries.

**The known:** COVID-19 has been diagnosed in over 4,000 Australians. Up until mid-March, most were from international travel, but now we are seeing a rise in locally acquired cases.

**The new:** This study uses a simple transmission dynamic model to demonstrate the difference between moderate changes to the reproduction number and forcing the reproduction number below one.

**The implications:** Lowering local transmission is becoming important in reducing the transmission of COVID-19. To maintain control of the epidemic, the focus should be on those in the community who do not regard themselves as at risk.

## Introduction

COVID-19 is a global problem, but the public health responses differ markedly between jurisdictions, particularly between countries. Australia is currently about two weeks behind the trajectory of similar high-resource nations such as the UK and the Netherlands ^1,2^. This delay has been achieved through our relative geographic isolation and strong response on international travel in the initial stages, beginning with travel restrictions to China (1st February) ^3^, then Iran (29^th^ Feb), South Korea (5^th^ March), and Italy (11^th^ March) ^4^. However, failure to identify emerging populations as at-risk in March (e.g. returning travelers from the US and the cruise ship the Ruby Princess) allowed a number of people to enter the country without testing for or awareness of infection with COVID-19. This finally led to an advisory against all non-essential travel for all countries on 20^th^ March 2020. Despite this strong stance, on 25^th^ March 2020, Australia surpassed China as the country with highest number of daily notified COVID-19 cases in the Western Pacific Region.

Now that sustained local transmission rivals importation as the main driver of new COVID-19 cases (including local transmission among people who are unaware that they are infected) there is a need for all Australians to change their behaviour. In this context, we need to examine just how stringent social distancing measures need to be to avoid most of our population becoming infected, and the associated loss of life.

A critical number in infectious diseases spread is the ***effective reproduction number, R***_***eff***_ (hereafter reproduction number) defined as the typical number of secondary infections that result from each person infected with the virus. Early in the Wuhan epidemic, this quantity sat at a value of around 2.7, but estimates have been wide-ranging ^5^. In settings similar to Australia, it has typically fallen in the range of 2.0 to 2.5, with a value of 2.4 being estimated consistently in models in the UK and Europe ^6^. Clearly if each person infects two to three others and the cycle repeats, exponential growth occurs in the initial phases. However, if we can reduce the reproduction number below one, then an epidemic will not occur. The daily growth rate is highly dependent on this quantity as well as the time between one generation of infections and the next, which has been estimated at 6.5 days ^6,7^, and is likely to be similar for all countries.

The effective reproduction number is at its highest when the virus is first introduced into a population and everyone is susceptible (at this point ***R***_***eff***_ is referred to as the ***basic reproduction number*** and is denoted by ***R***_***0***_). As the infection sweeps through the population, the proportion of susceptible individuals begins to decline and the effective reproduction number drops in direct relation to this proportion. Herd immunity occurs when a sufficient fraction of the population is immune to allow the effective reproduction number to fall below one. For this to occur for a virus with a basic reproduction number of *R*_*0*_ = 2.4, we need a little under 1 – 1 / *R*_*0*_ ≈ 60% of the population immune. However, 60% of Australia (or approximately 15 million people) acquiring immunity through uncontrolled natural infection may equate to tens or hundreds of thousands of deaths.

Enacting intervention strategies such as social distancing or vaccination can further reduce the effective reproduction number. However, unless these strategies are maintained – or a sufficient a fraction of the population develops immunity – higher values of *R*_eff_ may be restored and secondary epidemic waves experienced.

Infectious diseases models are being used more than in any previous epidemic to understand and tackle COVID-19. A recent study by Ferguson *et al* of Imperial College suggests that even by modifying the transmission rate by public health interventions that combine home isolation of suspect cases, home quarantine of those living in the same household as suspect cases, and social distancing of the elderly and others at most risk of severe disease (i.e., a mitigation strategy), the UK would still experience an epidemic which would overwhelm current health services many times over ^6^. Applying these results directly to Australia’s national or jurisdictional demography is similarly alarming ^8^.

Therefore a much expanded effort to control the epidemic has been advocated. Achieving a “suppression strategy”, which would bring the reproduction number below one, would require a combination of social distancing of the entire population, home isolation of identified cases and household quarantine of their family members as a minimum effort ^6^. This model was specified for the UK and US based on demography and detailed agent-based features, such as having specific country-level representation of transmission in households, schools, workplaces, and the wider community ^9^.

However, the problem with the suppression strategy is that herd immunity is not achieved and hence the measures taken need to be sustained or the epidemic will re-establish. The population dynamic results from Ferguson *et al* (although not the detailed impact of specific interventions) can be readily replicated by simple and transparent models, and adapted to the Australian setting, which we describe as follows.

## Methods

We developed a simple SEIR-type compartmental model (susceptible (S), exposed/incubation period (E), infectious (I) and removed(R)), Figure 1. The standard model is modified to allow for pre-symptomatic transmission during the incubation period (E_2_), a delay between the onset of symptoms and presentation to healthcare (I_1_), diagnosed disease (I_2_), hospitalisation (H), and ICU admission (ICU). In this simple model, compartments E_2_, I_1_ and I_2_ are infectious, while the H and ICU compartments are not. The H and ICU compartments represent people who have presented for care and make up around 5% of all infected people in the model. In reality these individuals would be isolated, such that we assume that the contribution of these groups to the force of infection is negligible. N, a fixed parameter not shown in Figure 1 is the total population:

**Figure 1.**
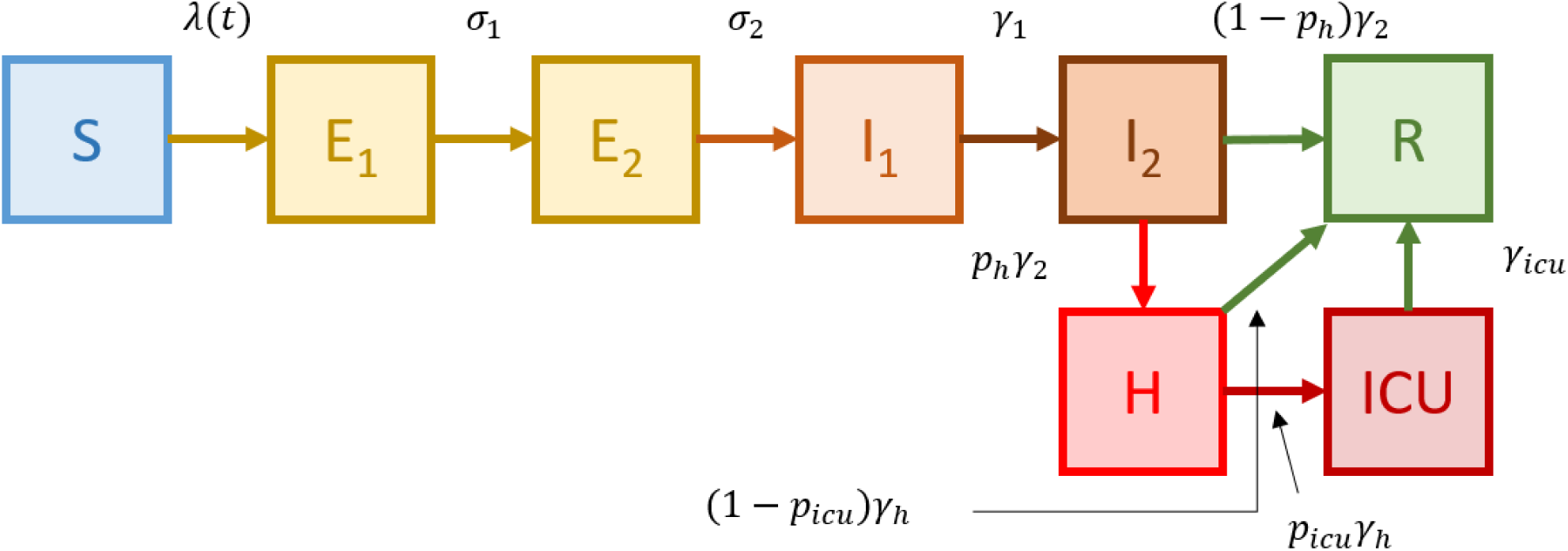
The modified SEIR model.

N=S+ E_1_ + E_2_ + I_1_ + I_2_ + H + ICU + R. The proportion of the population in each compartment is denoted using lower case, e.g. s=S/N.

The force of infection is therefore given by

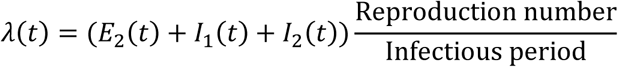

Where

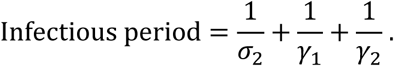

### Initial conditions

We begin each simulation on 25^th^ January assuming there had been no local cases in Australia prior to this date, with all individuals susceptible (*s(0)=1*). We then allow notified imported cases to enter the model into the I_2_ compartment on the day they were notified.

#### Calibration

We commenced with only reported imported cases in the model and the assumption of equivalent infectiousness to community-infected cases. The infectiousness of imported cases relative to the infectiousness of other cases was then varied until the modelled epidemic curve was similar to that seen in Australia. The calibrated infectiousness of known imports shown in Figure 2 was one sixth the infectiousness of a typical person who is locally infected.

**Figure 2.**
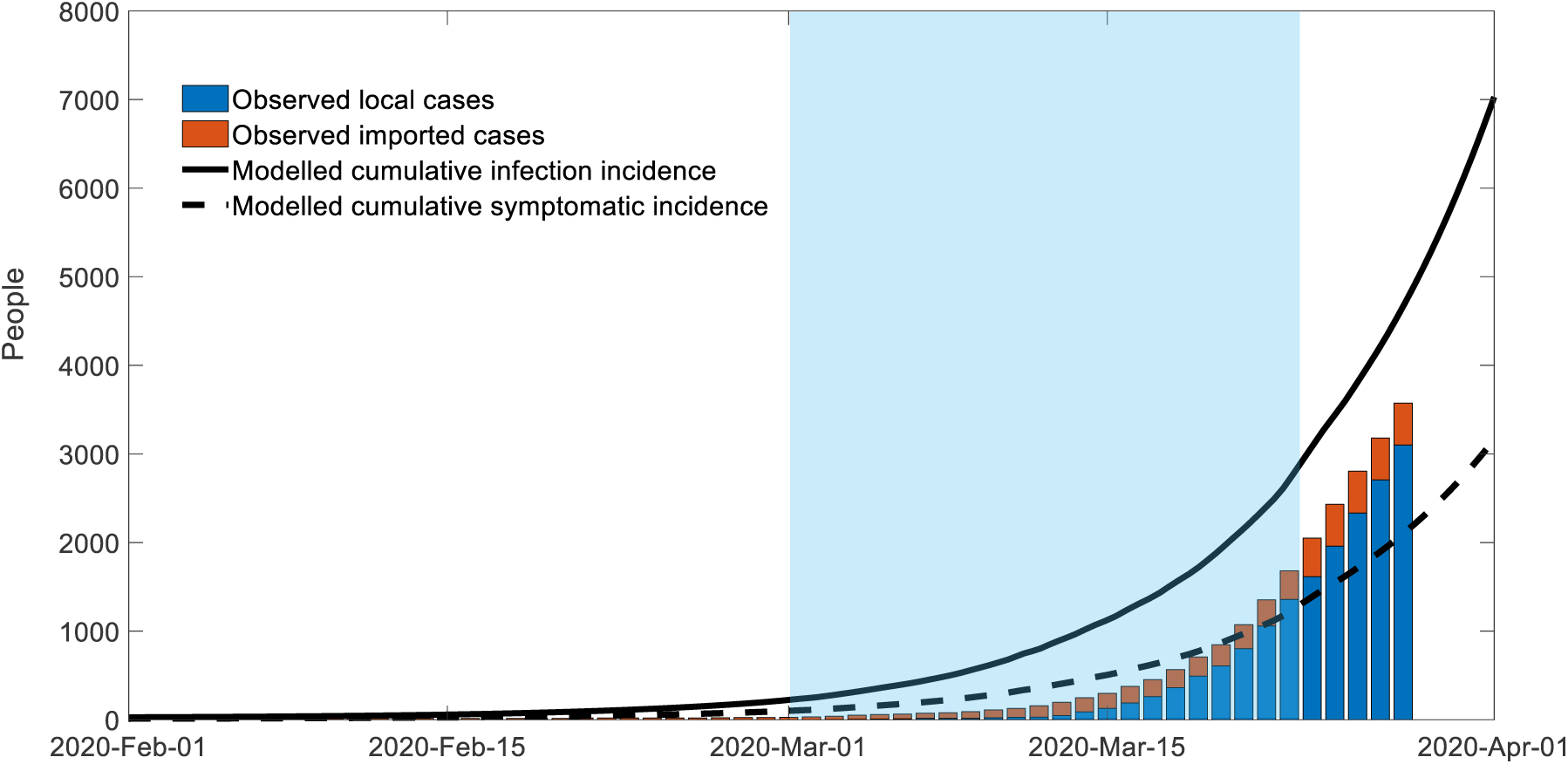
Stacked bar plot of cumulative notified imported cases (brown) and cumulative notified local cases (blue) in Australia from 1^st^ February until 25^th^ March. Calibrated model, with all infected people (solid line) and symptomatic infected people (dashed line). The blue background indicates the period of rapid change in testing regimens. Some cases were not yet defined as local or imported and these were assumed to be locally acquired.

In estimating the calibrated values, we additionally considered the rapid increase in testing that occurred from early March until 25^th^ March. During this time, Australia expanded its testing criteria, increasing the number of people tested compared with other settings ^13^ and finding a number of low symptomatic and asymptomatic cases, although exact approaches varied by jurisdiction. This effect was simulated by calibrating such that symptomatic infections (dashed line) begins above the observed cases (suggesting under-detection), but by 25^th^ March is below the observed cases, reflecting high testing rates including detection of asymptomatic cases.

#### Scenario analysis

To illustrate the spectrum of intervention strategies from baseline (no interventions) through mitigation to suppression, we consider four separate scenarios with different effective reproduction numbers outlined below:

#### Scenario 1: business as usual, *R*_*eff*_ = 2.4

This is examined by projecting forward the epidemic model with all parameters unchanged, including the reproduction which remains at 2.4.

#### Scenario 2: flatten and shift the curve, *R*_*eff*_ = 1.6

We consider the impact of reducing the reproduction number by one third. Conceptually, this means that for every three people infected, one does not pass on the disease, while the other two behave as in the business as usual scenario. Alternatively, it could be considered all infected individuals reduce their contacts by one third. Currently, it is too early to tell how effective transmission reduction strategies are; however, conceivably, intensive case and contact finding, with quarantining of those known to be contacts or at risk through travel, and isolating cases and their families, a “search, test and isolate” strategy could reduce onward transmission by one third. This would be similar to the response of Japan, which was initially highly effective at suppressing the outbreak, although in recent weeks has had an estimated reproduction ratio of between 1 and 2 ^10^.

#### Scenario 3: slow burn and maintain, *R*_*eff*_ = 1.17

Similar to scenario two, in the slow burn scenario we consider an exaggerated flattening process whereby the reproduction number is reduced to such an extent that the maximum ICU utilisation rate does not exceed the current national ICU capacity (approximately 2200 beds). For the baseline parameter values specified in Table 1 we find that this occurs for *R*_*eff*_ = 1.17.

**Table 1.** Parameters in the model

| Parameter | Value | Explanation |
| --- | --- | --- |
| **Reproduction number** | $R_{eff}$ | Typical number of secondary infections per infected person |
| **Business as usual** | 2.4 | Reproduction number in the absence of interventions |
| **Flatten the curve and achieve herd immunity** | 1.6 | Reproduction number under a combination of home isolation of suspect cases, home quarantine of those living in the same household as suspect cases, and social distancing of the elderly and others at most risk. Vigorous contact tracing and testing. Example countries: Japan <sup>10</sup> |
| **Further flatten the curve to allow very low and slow epidemic** | 1.17 | As above plus some level of population-wide social distancing, cancellation of large events and reduced unnecessary gatherings, work from home if possible. Example countries: Singapore <sup>10</sup> |
| **Contain and control** | 0.8 | Reproduction number under conditions of universal social distancing measures, population home quarantine and lock-down. Example settings: Hubei Province, South Korea <sup>10</sup> |
| **Duration of time in early incubation (uninfectious)** | $(\sigma_1)^{-1} = 3.6$ days | <sup>6</sup> |
| **Duration of time infectious prior to symptoms** | $(\sigma_2)^{-1} =$ half a day | <sup>6</sup> |
| **Early infectious period** | $(\gamma_1)^{-1} = 2$ days | Symptomatic period prior to detection |
| **Late infectious period** | $(\gamma_2)^{-1} = 5.68$ days | Remaining infectious period |
| **Duration of time in hospital** | $(\gamma_h)^{-1} = 8$ days | To match observations in Italy (personal correspondence described in Ferguson *et al* <sup>6</sup>) |
| **Duration of time in ICU** | $(\gamma_{icu})^{-1} = 10$ days | To match observations in Italy (personal correspondence described in Ferguson *et al* <sup>6</sup>) |
| **Initial conditions** | No local cases at start of epidemic 25<sup>th</sup> January 2020. | Imported cases as per Figure 2. Data sourced from www.COVID19data.com.au <sup>11</sup> |
| **Proportion of people hospitalised** | 4.4 % of cases<br>2.2% of infections | <sup>6</sup> |
| **Proportion of hospitalisations admitted to ICU** | 30% | <sup>6</sup> |
| **Per infectious person daily infectiousness** | $\beta = \frac{R_{eff}}{1/\sigma_2 + 1/\gamma_1 + 1/\gamma_2}$ | In this model, a simplifying assumption of equal infectiousness was used for all stages of infection until hospitalisation |
| **Force of infection** | $\beta s(t)^{\alpha} i(t)$ | |
| **Dissipation of infectiousness as the proportion of the s(t) reduces** | $\alpha = 1.18$ | Calibrated to Ferguson to allow an $R_{eff} = 2.4$ and a final size =81% <sup>6</sup> |
| **Probability of symptoms given infection** | 0.45 (45%) | <sup>6,12</sup> |
| **Infectiousness of known imports relative to local cases** | 0.16 | Fitted during calibration |

#### Scenario 4: flatten the curve the “Suppression” scenario, *R*_*eff*_ = 0.8

We examine the impact achievable if we reduce the reproduction number to below one. This would require a reduction by approximately two thirds; equivalent to two out of three people undergoing complete isolation for the duration of their infectious phase or all people reducing their contacts to around one third of contacts usually taking place. Measures taken would need to be similar to those in South Korea in March 2020 and Hubei since mid-February 2020. These regions have been able to achieve reproduction numbers below one ^10^.

## Results

The outcomes of having a reproduction number of 2.4 to 1.6 or 1.17 from 1^st^ April 2020 are shown in Figure 3.

**Figure 3.**
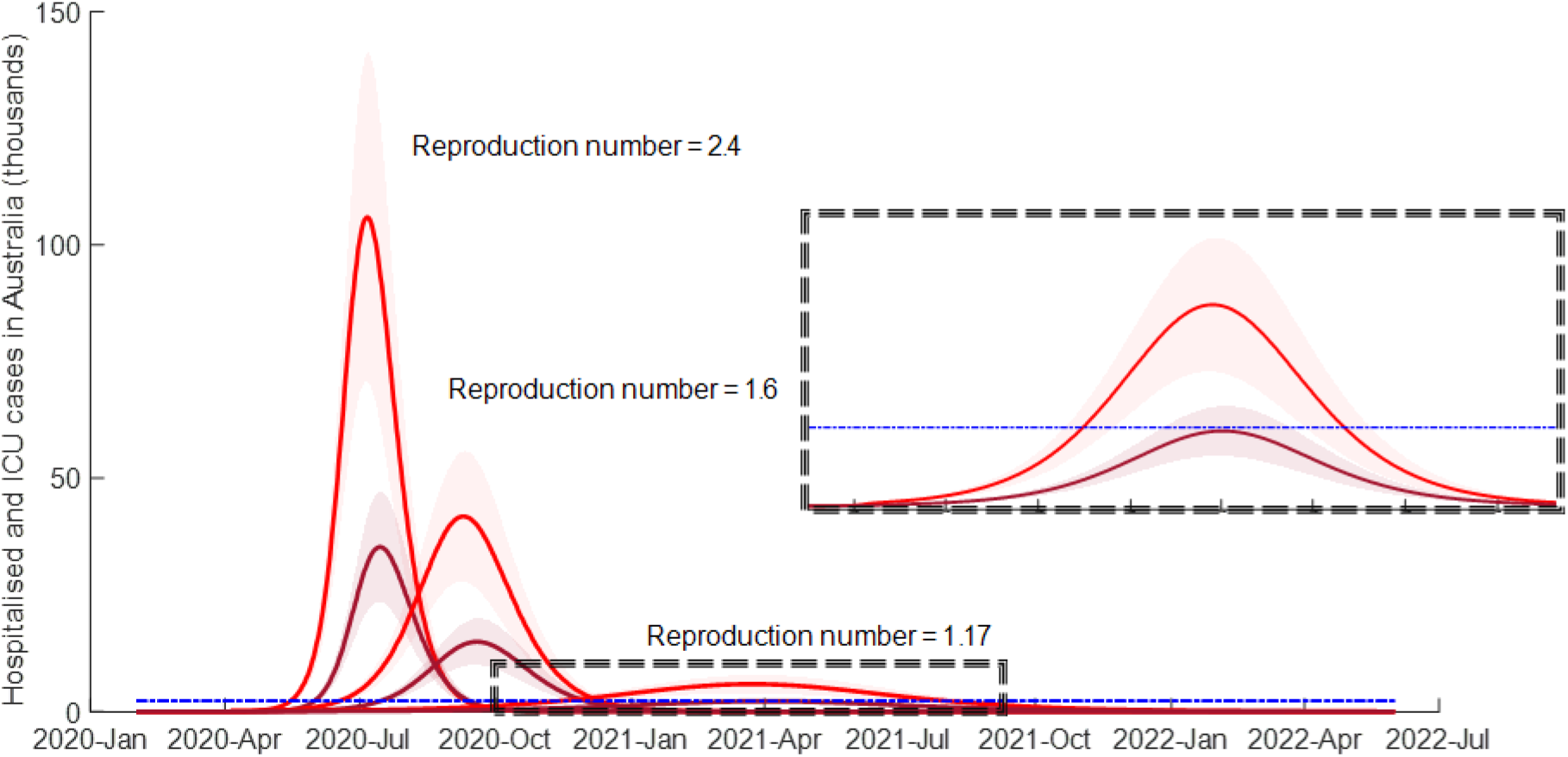
Modelled prevalence of people requiring hospitalisation (red lines) and ICU (maroon lines) for COVID-19. The shaded regions correspond to hospitalisation rates of 2% to 4% with the bold lines representing a 3% infection hospitalisation rate. This was modelled assuming reproduction numbers of 2.4, 1.6 and 1.17 as indicated. Australia’s ICU bed capacity is indicated by the blue dashed line.

### Scenario 1: business as usual, *R*_*eff*_ = 2.4

The modelled outcome of allowing COVID-19 to go unchecked is presented in Figure 2. Under these circumstances, we predict that at the epidemic’s peak 35,000 people will require critical care and around 105,000 will require hospitalisation. This would severely overwhelm hospitals and ICU facilities. The peak in transmission would occur in early July and the peak in hospital utilisation in mid-July, with 7% of the population symptomatic at this time. Around 81% of the population would be expected to be infected by the end of the epidemic.

### Scenario 2: flatten and shift the curve, *R*_*eff*_ = 1.6

We estimate that the epidemic peak of infectious cases would shift to October and peak ICU utilisation will shift to November, with a peak requirement for ICU beds of 15,000 and hospital beds of 42,000. At the peak, 3% of the population would be symptomatic. Around 57% of the population would be expected to be infected by the end of the epidemic.

### Scenario 3: slow burn, *R*_*eff*_ = 1.17

In Scenario 3 we examine a degree of infectiousness required to allow the epidemic to proceed at a slow enough pace to continue but not to exceed ICU capacity. This requires a reproduction ratio of no more than 1.17, and will lead to a peak utilisation of 2200 ICU beds and 5800 hospital beds. Only 0.4% of the population would be symptomatic at the peak, which is estimated to arrive in April 2021. Note that in this scenario the epidemic extends well into 2022. Approximately 24% of the population would be infected by the end of the epidemic.

### Scenario 4. Suppression, *R*_*eff*_ = 0.8

In Scenario 4, we assume a reproduction ratio of 0.8. This means that, on average, people transmit infection to less than one other person. This would be consistent with some transmission events and even a few links in a chain of transmission, but would not result in large ongoing clusters of transmission in the community.

In the suppression scenario, herd immunity is not achieved during the initial epidemic (see Figure 5). The result is that as soon as restrictions are lifted, secondary epidemic waves are almost inevitable. Therefore, successful elimination in this context requires several intermittent periods of strong interventions.

Figure 4 shows how intermittent suppression could translate in epidemic terms; the reproduction rate oscillating between 0.8 and 2.4 corresponding to periods with and without interventions, respectively. The hypothetical scenario shown in Figure 4 applies increased stringency measures when 1000 people are showing symptoms and withdraws these measures when fewer than 10 people are showing symptoms. In this scenario, the number of ICU beds required remains manageable and only around 0.5 % of the population become infected in these first three waves. Note the blue dashed line is at the same level as in Figure 3, with a very different scale.

**Figure 4.**
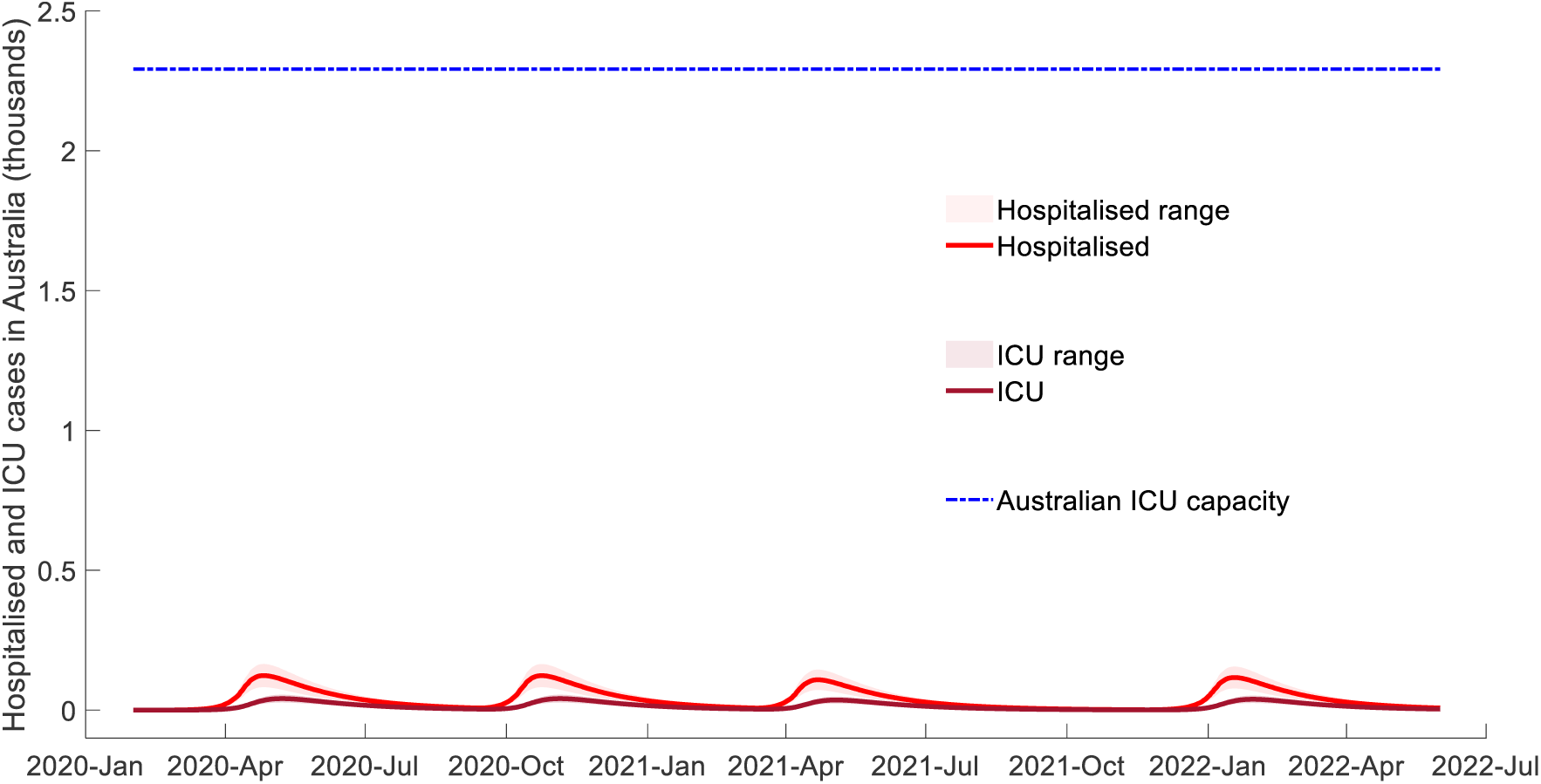
Impact of intermittent strategy: the prevalence of cases requiring hospitalisation under the intermittent R=2.4/R=0.8 strategy (red lines), number of COVID-19 cases requiring ICU admission (maroon lines).

### Herd Immunity

For a basic reproduction number of 2.4, the threshold proportion of people who must be resistant to infection (through natural immunity or vaccination) to forestall the epidemic is 1-1/R_0,_ or approximately 60%^1^. This equates to ∼ 15 million people in Australia, as shown by the green line in Figure 5. In general, if an epidemic is left unchecked, the number of incident infections begins to decrease when the population achieves herd immunity, and by the end of the epidemic, more of the population than is required to achieve herd immunity will have been infected (that is, some overshoot will occur). For example, for the baseline scenario with *R*_*eff*_ = 2.4 envisaged, rather than the required 60% herd immunity, the model estimates approximately 80% immunity at the end of the epidemic.

**Figure 5.**
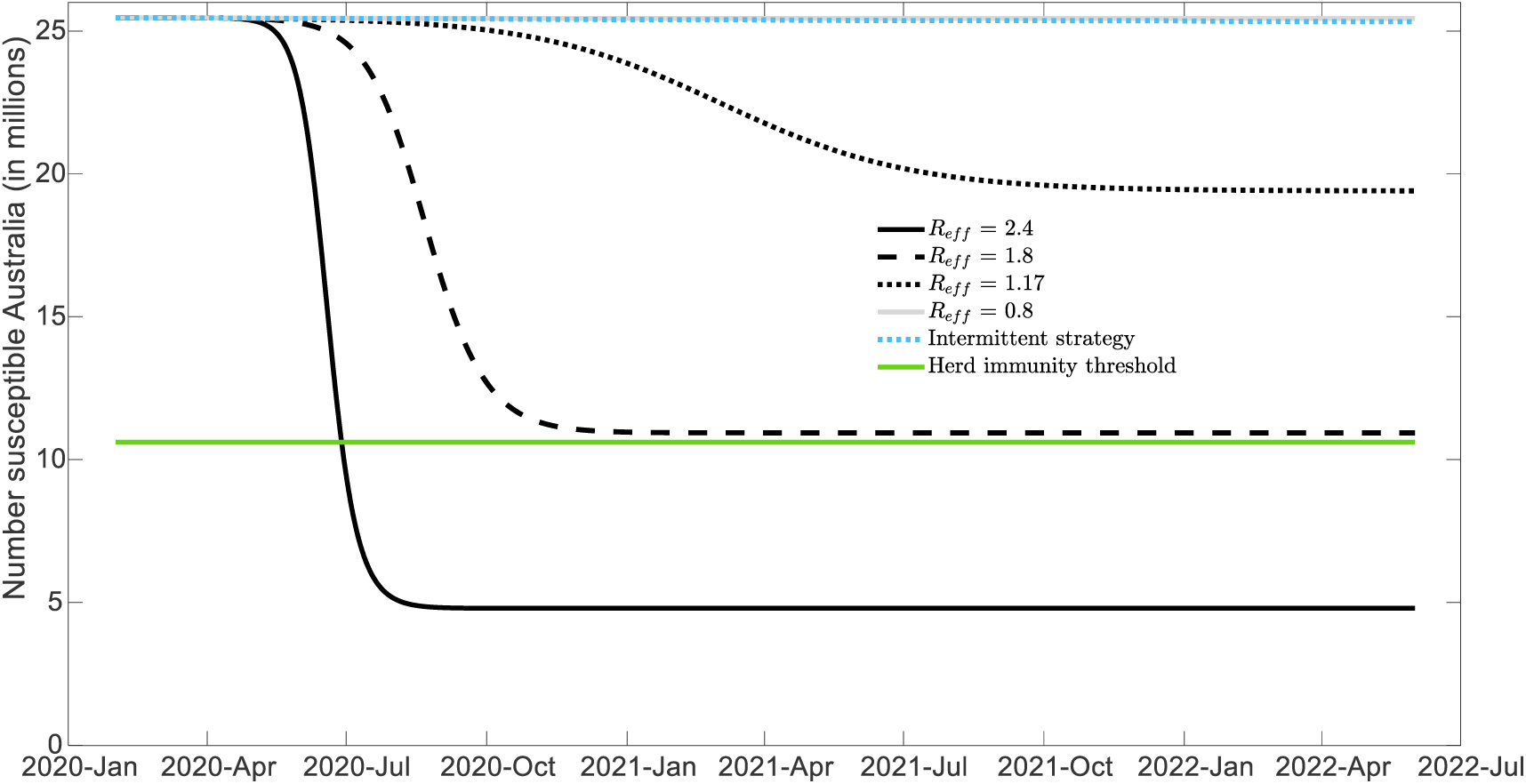
Herd immunity threshold (assuming R_0_ =2.4) in green compared to the expected number of people to remain susceptible in Australia depending on the strategy.

From Figure 5, we also observe that herd immunity is very close to being achieved in the first flattening scenario with *R*_*eff*_ = 1.6. In this case we would expect limited transmission following the first epidemic wave even after social distancing and other intervention measures are lifted. Conversely, in Scenarios 3 and 4 with *R*_*eff*_ = 1.17 and 0.8 respectively, the fraction of the population infected during the first wave would be insufficient to prevent future outbreaks occurring.

## Discussion

The purpose of this study was to provide a simple compartmental model to explain the impact of mitigation and suppression effects in a transparent way. We applied the concepts of mitigation and suppression to the Australian COVID-19 epidemic, calibrated to our current epidemic curve as at 27^th^ March 2020. Our results show that with a reproduction number of 2.4 and in the absence of a response to the epidemic, we can expect the majority of Australians to be infected by the end of winter 2020, with a peak expected in early July. Mitigation measures, as simulated in Scenario 2, dramatically reduce the rate of upswing of the epidemic curve, and delay its peak until early November. However, mitigation measures that only reduce the reproduction number to 1.6 have only a modest effect on the final number of people infected. Aiming for herd immunity and slowing the epidemic peak by 5 to 6 months will still overwhelm the health system (albeit by about 7-fold rather, than 16-fold) and the associated excess deaths would be unthinkable. We could do our best to protect the elderly from the illness to reduce deaths, but it is unlikely that we could avoid enormous numbers of fatalities even with moderate strategies in place that reduce the reproduction number to 1.6. On the other hand, slightly more stringent measures that reduce the reproduction number to 1.2 and maintain ICU case numbers below current capacity would extend the duration of the epidemic for more than two years, which seems an impractical strategy.

We must now turn our attention to how we may achieve suppression of COVID-19 transmission to enable the effective reproduction number to fall below one. We must look to countries and jurisdictions that have achieved this feat, such as South Korea since 2^nd^ March and Hubei since 20^th^ February, 2020. These two settings have taken very different approaches, with South Korea conducting high numbers of tests (around 40 tests for every confirmed case) ^14^, while Hubei has used travel restrictions and universal social distancing with home lockdown ^15^. Singapore and Japan have taken different paths to China, in relying primarily on test and manage tactics with cancellation of some major events, but continuation of work, schools and life ^15^. This is encouraging, but we need to watch carefully as the pandemic proceeds, as both countries are now experiencing gradual renewed growth in their epidemics, suggesting reproduction numbers above one – at least temporarily.

If cases of infection can be suppressed as has been reported for Hubei province, a series of follow-on questions must be posed. We need to consider which components of the intervention were most effective, and how long they can be maintained. Sero-surveys in the near future will provide important information of the number of undiagnosed cases; however, it seems implausible that even Hubei has had sufficient infection to provide herd immunity, which would require around 6 to 7 million infections in the Province (for a basic reproduction number of 2.4).

If we are lucky and control the first epidemic wave in Australia, our next question will be how to tackle the absence of herd immunity following suppression of the virus. Fundamentally, there are only three possibilities: 1) relax restrictions and allow an epidemic with similar characteristics to the first wave, 2) maintain suppression until vaccination is possible (or indefinitely), and 3) allow limited circulation to occur while protecting the vulnerable. Each of these approaches is associated with enormous costs. For the first, in the absence of population immunity, the burden of disease and loss of life would be similar to that predicted under our *R*_*eff*_ *> 1* scenarios. For the second, it remains unclear whether suppressive measures that can maintain *R*_*eff*_ *< 1* are compatible with a functioning society and economy, while the timing of the arrival of a novel vaccine remains similarly uncertain. The third would require segregation of society to isolate the elderly and at-risk from controlled transmission in younger groups, which would require unprecedented changes to our social structures and/or lifestyles. Ferguson *et al*. explored time out periods to allow financial recovery and re-instate these measures after a threshold is again reached ^6^. From an exclusively epidemiological standpoint, there is no particular reason to conclude that this would minimise the impact of interventions, but could be considered if pulsed relaxations were beneficial from a social/economic perspective, as suggested by the authors.

Another approach, is to take the foot off the brake slowly and watch and wait. This could involve restoring those parts of society that are most crucial and least vulnerable – such as schools and some workplaces, followed by domestic travel and travel to other countries performing equally well in containing the virus. Knowledge of social mixing patterns and the marked differences in vulnerability with age could be used to explore approaches for relaxing contact restrictions in such a way as to minimise harm but permit some circulation of the virus. Each of the possible responses are associated with major potential adverse impacts for our country, and our choices should be guided by the values of our society. In the short term, we strongly support contact restrictions to gain temporary control and begin longer term planning from a position of stability.

Australia is now on the brink. The number of local cases may soon exceed our ability to test and trace, although Australia has enviable testing capacity, with over 181,000 tests already performed ^16^. Therefore universal measures will have to be taken to slow community transmission, early signs show cause for optimism with a slight slowing of incident cases at the end of March. The next two weeks will be critical to Australia’s ability to suppress COVID-19, and a modest strategy or a strategy aimed only at those at highest risk will fail to prevent a large epidemic.

## Data Availability

Data and code used in this manuscript are provided in the supporting material

## Appendix A

**Table A1.** Calculation of expected infection fatality rate in Australia, based on Australian age distribution and age-specific estimated infection fatality rate in Wuhan Province, China.

| **Age Group** | **Proportion of Australian population<sup>17</sup> (%)** | **IFR<sup>12</sup>** | **Contribution to Australian IFR** |
| --- | --- | --- | --- |
| 0–4 | 6.4% | 0.0016% | 0.00% |
| 5–9 | 6.6% | 0.0016% | 0.00% |
| 10–14 | 6.4% | 0.0070% | 0.00% |
| 15–19 | 6.1% | 0.0070% | 0.00% |
| 20–24 | 7.2% | 0.031% | 0.00% |
| 25–29 | 7.6% | 0.031% | 0.00% |
| 30–34 | 7.4% | 0.084% | 0.01% |
| 35–39 | 7.0% | 0.084% | 0.01% |
| 40–44 | 6.3% | 0.16% | 0.01% |
| 45–49 | 6.6% | 0.16% | 0.01% |
| 50–54 | 6.0% | 0.60% | 0.04% |
| 55–59 | 6.0% | 0.60% | 0.04% |
| 60–64 | 5.4% | 1.9% | 0.10% |
| 65–69 | 4.7% | 1.9% | 0.09% |
| 70–74 | 4.1% | 4.3% | 0.18% |
| 75–79 | 2.8% | 4.3% | 0.12% |
| 80–84 | 1.8% | 7.8% | 0.14% |
| 85 and over | 1.6% | 7.8% | 0.12% |
| **All ages** | **100.0** |  | **0.86%** |

**Table A2.** Calculation of expected case hospitalisation proportion per symptomatic case in Australia, based on Australian age distribution and age-specific estimated case-hospitalisation proportion in Wuhan Province, China.

| Age Group | Proportion of Australian population (%) | Case Hospitalisation proportion <sup>12</sup> | Contribution to case hospitalisation proportion |
| --- | --- | --- | --- |
| 0–4 | 6.4% | 0.1% | 0.0% |
| 5–9 | 6.6% | 0.1% | 0.0% |
| 10–14 | 6.4% | 0.3% | 0.0% |
| 15–19 | 6.1% | 0.3% | 0.0% |
| 20–24 | 7.2% | 1.2% | 0.1% |
| 25–29 | 7.6% | 1.2% | 0.1% |
| 30–34 | 7.4% | 3.2% | 0.2% |
| 35–39 | 7.0% | 3.2% | 0.2% |
| 40–44 | 6.3% | 4.9% | 0.3% |
| 45–49 | 6.6% | 4.9% | 0.3% |
| 50–54 | 6.0% | 10.2% | 0.6% |
| 55–59 | 6.0% | 10.2% | 0.6% |
| 60–64 | 5.4% | 16.6% | 0.9% |
| 65–69 | 4.7% | 16.6% | 0.8% |
| 70–74 | 4.1% | 24.3% | 1.0% |
| 75–79 | 2.8% | 24.3% | 0.7% |
| 80–84 | 1.8% | 27.3% | 0.5% |
| 85 and over | 1.6% | 27.3% | 0.4% |
| All ages | 100.0% |  | **6.8%** |

Proportion of cases that are symptomatic is estimated at 45% <sup>6</sup>, therefore anticipated infection hospitalization proportion are $0.45 \times 6.8\% = 3.08\%$ . We examined proportions of 2, 3 and 4% in the study.

## Footnotes

1 This is a general finding for the most homogenous mixing model. Readers may have noticed that we have generalised this slightly, such that the herd immunity threshold is 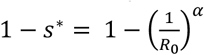. Because α is slightly above one, this leads to a slightly higher (more optimistic) herd immunity.

## References

1. Burn-Murdoch J. An interactive visualization of the exponential spread of COVID-19. 2020. http://91-divoc.com/pages/covid-visualization/ (accessed 27-Mar-2020.

2. JHU. Coronavirus COVID-19 Global Cases by the Center for Systems Science and Engineering (CSSE) at Johns Hopkins University (JHU). 2020. https://gisanddata.maps.arcgis.com/apps/opsdashboard/index.html#/bda7594740fd40299423467b48e9ecf6 (accessed 25-Mar 2020).

3. 50. ABC. Australians told not to travel to mainland China due to coronavirus threat, border restrictions tightened considerably. 2 Feb 2020. 2020. https://www.abc.net.au/news/2020-02-01/australians-told-not-to-travel-to-china-due-to-coronavirus/11920742 (accessed 27 Mar 2020).

4. 51. VOA. Australia Extends Virus Travel Ban to Italy. 2020. https://www.voanews.com/science-health/coronavirus-outbreak/australia-extends-virus-travel-ban-italy (accessed 12/03/2020 2020).

5. McBryde E. The value of early transmission dynamic studies in emerging infectious diseases. The Lancet Infectious Diseases 2020.

6. Ferguson NM, Laydon D, Nedjati-Gilani G, et al. Impact of non-pharmaceutical interventions (NPIs) to reduce COVID-19 mortality and healthcare demand. 2020.

7. Wu JT, Leung K, Bushman M, et al. Estimating clinical severity of COVID-19 from the transmission dynamics in Wuhan, China. Nature Medicine 2020: 1–5.

8. Fox GJ, Trauer JM, McBryde ES. Modelling the impact of COVID-19 upon intensive care services in New South Wales Medical Journal of Australia 2020; in press: accepted 27th March 2020.

9. Ferguson NM, Cummings DAT, Cauchemez S, et al. Strategies for containing an emerging influenza pandemic in Southeast Asia. Nature 2005; 437(7056): 209–14.

10. Abbott S, Hellewell J, Munday JD, et al. Temporal variation in transmission during the COVID-19 outbreak. 2020. https://cmmid.github.io/topics/covid19/current-patterns-transmission/global-time-varying-transmission.html (accessed 27/03/2020.

11. Bal T, Selladoray A, de Graaf R, Kaushik N. Coronavirus (COVID-19) in Australia. 2020. https://www.covid19data.com.au/ (accessed 27 Mrach.

12. Verity R, Okell LC, Dorigatti I, et al. Estimates of the severity of COVID-19 disease. medRxiv 2020: 2020.03.09.20033357.

13. Ortiz-Ospina E, Hasell J. How many tests for COVID-19 are being performed around the world? 2020. https://ourworldindata.org/covid-testing (accessed 25 Mar 2020).

14. Won S. COVID-19 confirmed, recovered, and test cases South Korea 2020: Case development trend. 2020. https://www.statista.com/statistics/1095848/south-korea-confirmed-and-suspected-coronavirus-cases/.

15. Cyranoski D. What China’s coronavirus response can teach the rest of the world. Nature 2020; 579(7800): 479–80.

16. WAnews. ‘One in every 220 tested’: How WA’s COVID-19 numbers compare to other states. 2020 (accessed 27-Mar 2020).

17. ABS. Australian Age Distribution Table. 2020. https://www.abs.gov.au/AUSSTATS/abs@.nsf/DetailsPage/3101.0Jun%202019?OpenDocument.

